# Relationship between mental health and unmet need for contraception and method type among women living with HIV in Kenya

**DOI:** 10.64898/2026.09.10.26362702

**Authors:** Agnes Karingo Karume, Aparna Seth, Nancy Ngumbau, Celestine Atieno, June Moraa, Kristin Beima-Sofie, Barbra A. Richardson, Jennifer A. Unger, Amritha Bhat, John Kinuthia, Alison L. Drake

## Abstract

Contraception is important for preventing unintended pregnancies and reducing maternal and infant mortality. Among women living with HIV (WLHIV), it is also central to preventing vertical HIV transmission. Yet unmet need for contraception remains high among WLHIV in sub-Saharan Africa. Mental health symptoms may contribute to unmet need, but evidence among WLHIV remains limited. We examined the relationship between depression and anxiety and unmet need for contraception.

We used baseline survey data from Kenyan WLHIV receiving routine HIV care participating in a cluster randomized clinical trial evaluating a reproductive health counselling intervention at 10 HIV clinics. Women who were fecund, did not desire a pregnancy within two years, and not using a modern method of contraception were considered to have unmet need for contraception and were eligible for this analysis. Surveys captured data on depression, anxiety, fertility intentions and contraceptive use. We constructed separate generalized linear models to assess the relationship between depression and/or anxiety and unmet need for contraception.

Among 2,334 women in the analysis, the median age was 34; 9% (n=221) reported symptoms of at least mild depression, 9% (n=200) anxiety, 13% (n=305) either condition, and 5% (n=116) both. Overall, 84% (n=1,952) used a modern contraceptive method, 16% (n=378) had an unmet need for contraception, and 41% (n=784) used LARC. In univariate analysis, unmet need for contraception was 43% higher among women with either at least mild depression or at least mild anxiety (Prevalence Ratio [PR]=1.43, 95% Confidence Interval [CI]: 1.22-1.67); results were similar in the adjusted model (adjusted PR [aPR]=1.30, 95% CI: 1.08-1.58). No relationships were detected between anxiety or depression alone and unmet need.

WLHIV with either depression or anxiety had higher unmet need which suggests that mental health conditions may pose barriers to contraceptive use and may need additional support to achieve their reproductive health goals. Additional attention to mental health needs during contraceptive counselling may support informed contraceptive decision-making.

## INTRODUCTION

Contraception is an important strategy to help women and girls prevent unintended pregnancies and reduce maternal and infant mortality(1)(2). For women living with HIV (WLHIV), contraception is also a cornerstone of prevention of vertical HIV transmission. Yet, in sub-Saharan Africa (SSA) where the HIV epidemic is concentrated, unmet need for contraception is high(3)(4). In Kenya, 15% of WLHIV have unmet need for contraception(5)(6) and 35% report their prior pregnancy was unintended(5). In addition, WLHIV who have unintended pregnancies are more likely to transmit HIV to infants, and have higher risks of maternal and infant mortality, than women who report their pregnancies were intended(7).

High unmet need among WLHIV has been attributed to factors such as stigma, limited integration of family planning (FP) services in HIV care, fear of partner disclosure, and socioeconomic vulnerabilities(5)(8)(9). Disproportionately higher rates of depression and anxiety among WLHIV may further contribute to elevated unmet need(10). For WLWH, contraceptive use may be impacted by depressive symptoms, via lack of self-efficacy or motivation related to negative attitudes or beliefs about their own reproductive health or fertility(11). In addition, women who experience depressive symptoms may find it difficult to proactively seek contraception or talk with a provider about contraceptive methods, lacking confidence or motivation to communicate their needs or initiate conversations with providers(12)(13). Similarly, anxiety may heighten fears about side effects attributed to contraceptive use, such as bleeding, mood changes and weight gain, which could impact their feelings towards using any contraceptive method(11)(13). Furthermore, anxiety may impair decision-making ability necessary to select a method and use it consistently(13). Finally, long-acting reversible contraception (LARC; implants and intrauterine contraceptive devices [IUCDs]), which require provider involvement for initiation and removal, may be less desirable for women who experience depression and/or anxiety(14). In Kenya, nearly one-quarter of WLHIV use LARC, the vast majority of which are implants(5)(6).

Studies conducted in the US have shown higher contraceptive non-use and discontinuation among women with depression and anxiety than those without depression or anxiety(12)(13)(15). Prior studies suggest young women with depression are more likely to use less effective methods such as oral contraception, condoms and withdrawal compared to LARC(13)(15); potentially because short-acting methods are easier to initiate than LARC, or they afford more control to continue or discontinue contraception due to the ability to use independently without provider assistance(16)(17). Strength of feelings about pregnancy prevention and fears of invasive procedures required for LARC may also differ between women who have depression or anxiety compared to those without these conditions. In a small retrospective study conducted in the U.S., there was a borderline association between having a mental health diagnosis and LARC removal within a year(18). However, the relationships between depression and anxiety and contraceptive use have not been well characterized in sub-Saharan Africa, nor among WLHIV, who may face unique or additional challenges in method use. We examined these relationships among Kenyan WLHIV receiving routine HIV care.

## METHODS

### Study design

We utilized baseline data from a cross-sectional survey administered to WLHIV enrolled in a cluster randomized controlled trial (cRCT) of a digital reproductive health counseling intervention, a tablet-based counseling tool delivered at enrollment and follow-up text communication with a study nurse over two-years, versus the standard of care(19).

### Site and population

The study was conducted at 10 HIV clinics located in four counties in Kenya: Kisumu, Homa Bay, Siaya and Nairobi; five clinics were randomized to intervention and five to control.

The cRCT enrolled 3298 women. To be eligible for the cRCT, women had to be living with HIV, of reproductive age (18-45; 14-17 if emancipated minors); have daily access to a mobile phone (own phone or shared) with a Safaricom SIM; plan to receive HIV care at the enrollment facility for 2 years; speak English, Kiswahili, or Luo; and be literate or comfortable with someone reading study SMS to them. Pregnant women were ineligible for enrollment. In addition, WLHIV were excluded from this analysis if they known to be infecund or intended to become pregnant within the next two years.

### Data collection procedures and measures

WLHIV attending routine HIV care at the HIV clinics located at each site were recruited to participate in the cRCT by study nurses, invited to participate if they met eligibility criteria, and provided written informed consent. Baseline surveys were administered to WLHIV after obtaining consent for study participation. At enrollment, study staff administered surveys on depression, anxiety, and stigma. Participants used a tablet to self-administer a FP survey, which assessed fertility intentions and contraceptive use. All surveys were administered in a private area within each facility in English or in a local language (Kiswahili or Dholuo) as preferred.

Depression was assessed using the Patient Health Questionnaire-9 (PHQ-9)(20), a 9-item scale ( range 0-27), with scores of 5, 10, 15, and 20 representing thresholds for mild, moderate, moderately severe, and severe depression, respectively. Anxiety was assessed using Generalized Anxiety Disorder-7 (GAD-7) scale (21), a 7-item rated on a 4-point (range 0-21) and scores of 5, 10, and 15 indicating mild, moderate and severe anxiety, respectively (21).

HIV-related stigma was assessed using the short version of the Berger HIV stigma scale (HSS12(22) (range12-48) with higher scores designating a greater perceived HIV-related stigma. The World Health Organization Violence Against Women (WHO-VAW) scale(23)(24) was used to assess intimate partner or gender-based violence with an affirmative response classified as intimate partner violence.

### Definitions and Statistical analysis

Modern contraceptive methods include LARC (implants and intrauterine contraceptive devices), short-acting contraceptive methods (condoms, pills, injectables, vaginal ring, lactation amenorrhea [LAM], standard day methods, emergency contraception) and permanent methods (tubal ligation, vasectomy). Unmet need for contraception was defined as the percentage of women who were fecund, had no desire for pregnancy in the next two years, and were not using modern contraception(25)(26).

The primary exposures were having at least mild depressive symptoms (PHQ-9 ≥5), having at least mild anxiety (GAD-7 ≥5), or having either depressive or anxiety symptoms. Sensitivity analyses were conducted to evaluate relationships with moderate depression and/or anxiety. Separate univariable generalized linear models (GLMs) with a log-link function and robust standard errors were constructed to examine the relationship between depression, anxiety, or both, and unmet need for contraception and separate models for LARC versus short acting. These models are appropriate for non-rare binary outcomes (27)(28). Variables significant at p≤0.1 in univariable analysis were included in multivariable models, along with the primary exposure. All models included facility as a fixed effect, with robust standard errors clustered by facility. There was 80% power at an α=0.05 to detect at least a 1.39-fold higher risk of unmet need for contraception among women with at least mild depressive symptoms. R studio version 4.2.2 was used for all analyses.

### Ethical Consideration

The study procedures were approved by the University of Washington Human Subjects Division (STUDY00013136), Kenyatta National Hospital-University of Nairobi Ethics Committee, (KNH-UoN ERC #P162/03/2022), and the National Commission for Science, Technology and Innovation (NACOSTI). All participants provided written informed consent before participation.

## RESULTS

### Demographic and clinical characteristics

Between September 2022 and June 2024, 4,129 women were screened, 3486 (84%) were eligible, 176 (4%) declined, and 3,298 were enrolled. Of these, 2,334 (71%) fecund women reported no intention to become pregnant within the next two years and were included in this analysis (Figure 1).

**Figure 1:**
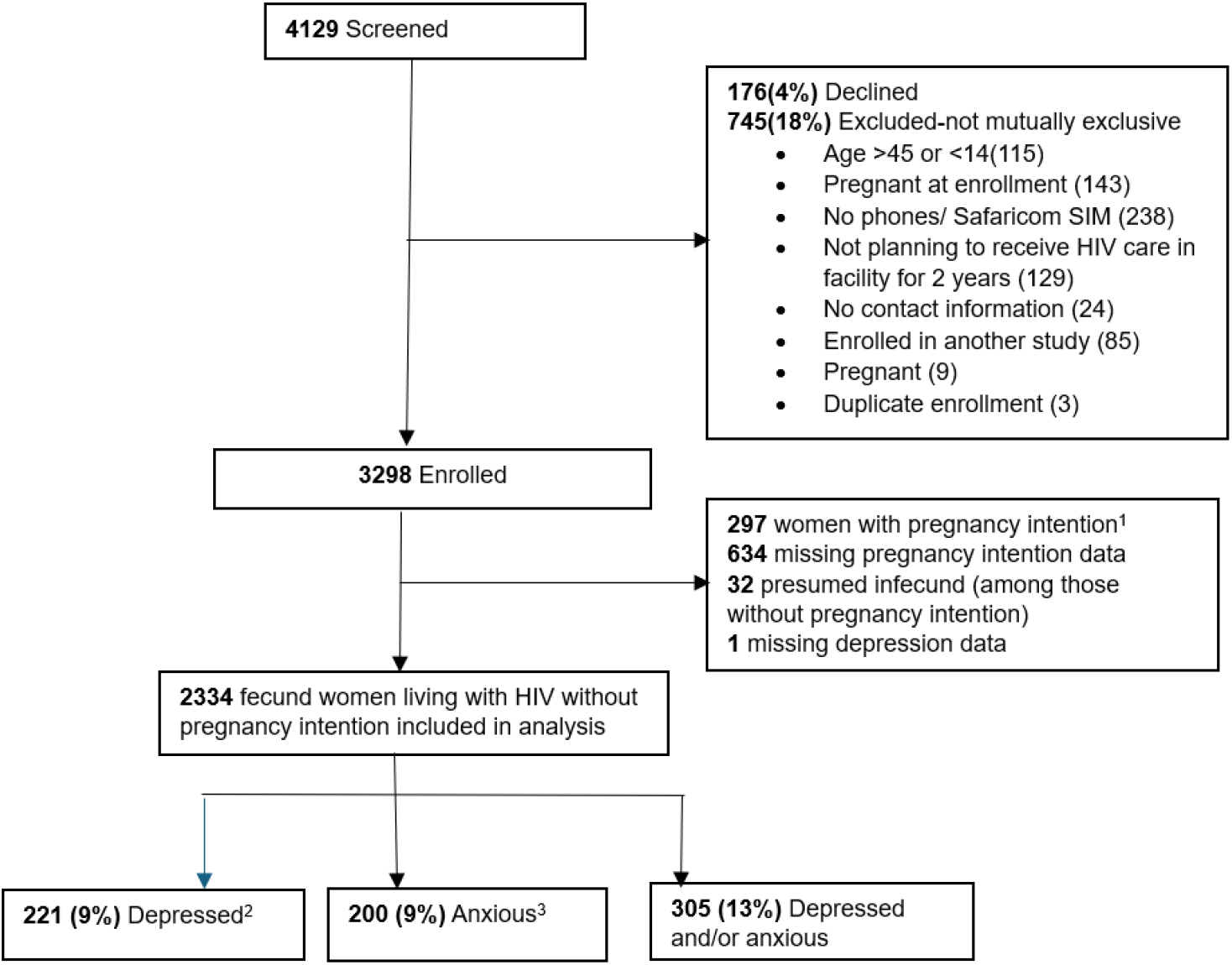
Study flowchart of Kenyan women living with HIV enrolled. ^1^Pregnancy intention – want a/another pregnancy within 2 years. ^2^Depressed - Patient Health Questionnaire 9 (PHQ-9) score ≥5; at least mild depression ^3^Anxious - Generalized anxiety disorder 7 (GAD-7) score ≥5; at least mild anxiety ^*^Depression and anxiety categories are not mutually exclusive

The median age was 34 years (interquartile range [IQR]:29-39) and most (64%, n=1,485) women were married or cohabiting (Table 1). The median gravidity was 3 (IQR 2-4). Nearly all women were on antiretroviral therapy (ART) (99%) and median time since HIV diagnosis was 8 years (IQR 4-12). The majority (70%, n=1,595) disclosed their HIV status to their partner. Overall, 79% (n=1,843) reported experiencing stigma, and 5% (86) experienced intimate partner violence. Based on self-reported symptoms, 9% (221) of WLHIV had at least mild depressive symptoms, 9% (200) had mild anxiety symptoms, 13% (305) had either depression or anxiety, and 5% (116) had both.

**Table 1:** Characteristics of Kenyan women living with HIV by depression and anxiety status.

| Characteristic | N | Overall<br>N= 2,334<br>n (%); Median (IQR) | Depressed<sup>1</sup><br>N = 221<br>n (%); Median (IQR) | Not depressed<br>N = 2,113<br>n (%); Median (IQR) | Anxiety<sup>2</sup><br>N=200<br>n (%); median (IQR) | No anxiety<br>N=2,134<br>n (%); median (IQR) |
| --- | --- | --- | --- | --- | --- | --- |
| **Socio-demographic** |  |  |  |  |  |  |
| Age (years) | 2,334 | 34 (29, 39) | 36 (30, 39) | 34 (29, 39) | 36 (31, 40) | 34 (29, 39) |
| At least secondary education | 2,334 | 1,078 (46%) | 91 (41%) | 987 (47%) | 80 (40%) | 998 (47%) |
| Married/cohabiting | 2,332 | 1,485 (64%) | 124 (57%) | 1,361 (64%) | 117 (59%) | 1,368 (64%) |
| Ever been pregnant | 2,334 | 2,269 (97%) | 218 (99%) | 2,051 (97%) | 199 (100%) | 2,070 (97%) |
| Gravidity (if ever pregnant) | 2,269 | 3 (2,4) | 4 (2,5) | 3 (2,4) | 4 (2,5) | 3 (2,4) |
| **HIV and partner characteristics** |  |  |  |  |  |  |
| Time since HIV diagnosis in years | 2,311 | 8 (4,12) | 9 (5,13) | 8 (4,12) | 9 (5,13) | 8 (4,12) |
| On ART | 2,334 | 2,315 (99%) | 220 (100%) | 2,095 (99%) | 196 (98%) | 2,119 (99%) |
| Have a regular partner | 2,326 | 1,727 (74%) | 150 (68%) | 1,577 (75%) | 135 (68%) | 1,592 (75%) |
| Partner HIV status | 1,727 |  |  |  |  |  |
| Positive |  | 949 (55%) | 87 (58%) | 862 (55%) | 77 (57%) | 872 (55%) |
| Negative |  | 565 (33%) | 45 (30%) | 520 (33%) | 42 (31%) | 523 (33%) |
| Don't know |  | 213 (12%) | 18 (12%) | 195 (12%) | 16 (12%) | 197 (12%) |
| Disclosed HIV status to partner | 2,287 | 1,595 (70%) | 144 (66%) | 1,451 (70%) | 127 (64%) | 1,468 (70%) |
| Experienced intimate partner violence (with partner)<sup>3</sup> | 1,715 | 86 (5%) | 34 (23%) | 52 (3%) | 27 (20%) | 59 (4%) |
| Experienced stigma<sup>4</sup> | 2,333 | 1,843 (79%) | 207 (94%) | 1,636 (77%) | 194 (97%) | 1,649 (77%) |
| **Contraception and sexual history** |  |  |  |  |  |  |
| Currently sexually active | 2,321 | 1,728 (74%) | 151 (70%) | 1,577 (75%) | 134 (68%) | 1,594 (75%) |
| Using modern contraception | 2,330 | 1,952 (84%) | 172 (78%) | 1,780 (84%) | 156 (78%) | 1,796 (84%) |
| Dual method use<sup>5</sup> | 1,952 | 369 (19%) | 36 (21%) | 333 (19%) | 22 (14%) | 347 (19%) |
| LARC use | 1,952 | 784 (40%) | 63 (37%) | 721 (41%) | 61 (39%) | 723 (40%) |
| Ever discussed contraception with provider since HIV diagnosis | 2,322 | 1,649 (71%) | 170 (78%) | 1,479 (70%) | 150 (76%) | 1,499 (71%) |
| Unmet need<sup>6</sup> | 2,330 | 378 (16%) | 49 (22%) | 329 (16%) | 44 (22%) | 334 (16%) |
| Intent to use contraception (among non-users) | 266 | 160 (60%) | 19 (68%) | 141 (59%) | 15 (63%) | 145 (60%) |

<sup>1</sup> Patient Health Questionnaire 9 (PHQ-9) score $\geq 5$ ; at least mild depression<sup>2</sup> Generalized anxiety disorder 7 (GAD-7) score $\geq 5$ ; at least mild anxiety<sup>3</sup> World Health Organization Violence Against Women score $\geq 1$ <sup>4</sup> Berger HIV Stigma Scale score $\geq 1$ <sup>5</sup> Dual method use: using condoms and another modern contraception method<sup>6</sup> Unmet need: Women who are fecund and do not want a/another child or do not desire pregnancy in the next two years, and are not using any modern method of contraception
ART=antiretroviral therapy; LARC=long-acting reversible contraception; IUCD (intra-uterine contraceptive device) and implant

### Contraceptive use

Overall, 84% (n=1,952) used modern contraception, 2% (n=48) used non-modern methods and 14% (n=330) did not use a method. The most frequently used methods of modern contraception were implants (37%, n=716) and injectables (36%, n=693), followed by male condoms (32%, n=633), oral contraception (8%, n=150) and IUCDs (4%, n=70) (Figure 2). Among modern contraceptive users, 19% (n=369) used dual methods.

**Figure 2:**
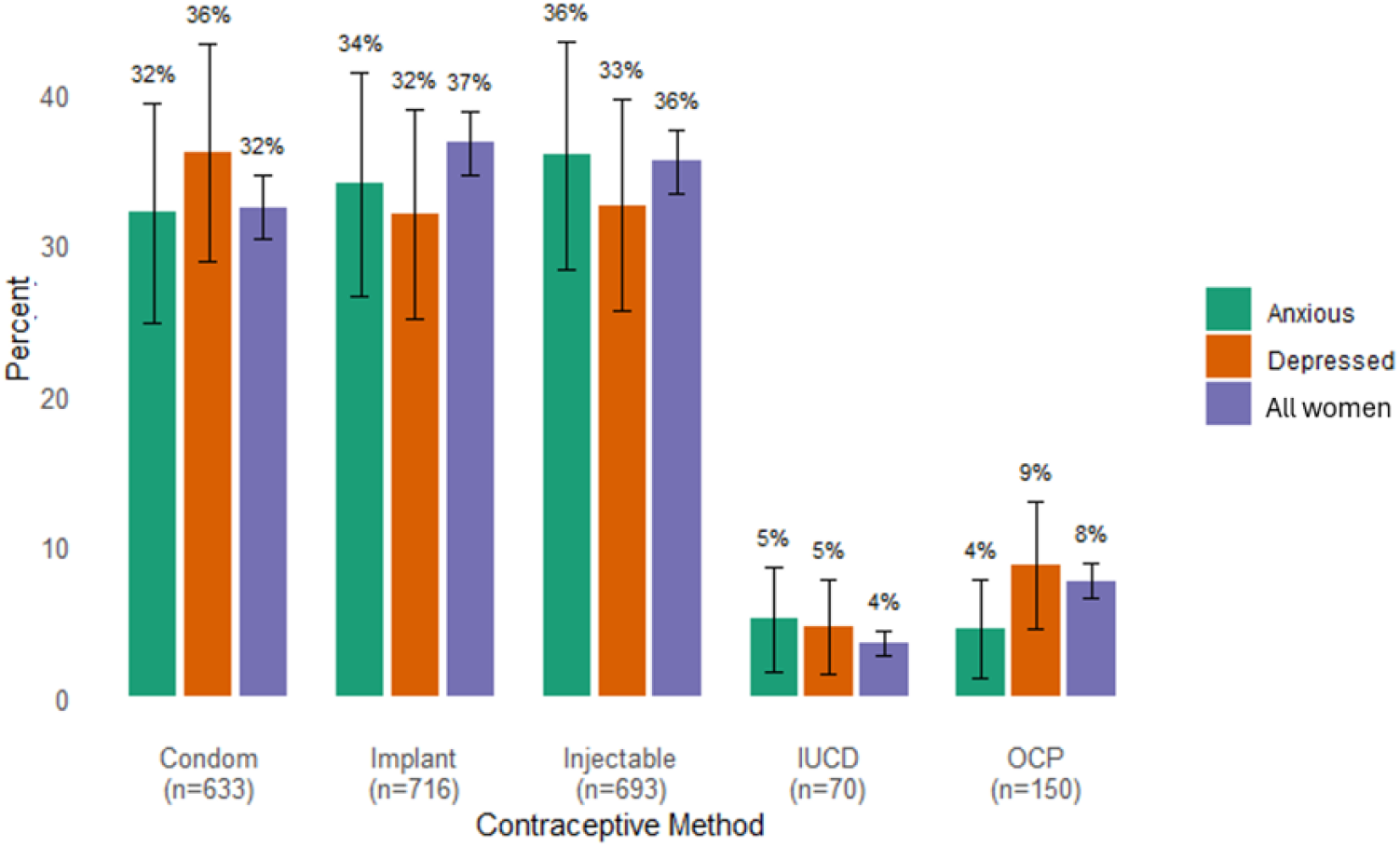
Contraceptive method mix excluding permanent methods among women living with HIV using modern contraception by anxiety or depression (n=1,952). IUCD - Intrauterine Contraceptive Device; OCP – Oral Contraceptive Pill Depressed - Patient Health Questionnaire 9 (PHQ-9) score ≥5; at least mild depression Anxious - Generalized anxiety disorder 7 (GAD-7) score ≥5; at least mild anxiety

### Unmet need for contraception

Overall, 16% (n=378) had an unmet need for contraception. In univariate analyses, unmet need was significantly higher among women with at least mild depressive symptoms (Prevalence Ratio [PR]:1.42, 95% Confidence Interval [CI]:1.20-1.68, p≤0.01), with symptoms of at least mild anxiety (PR 1.40, 95% CI 1.16, 1.70, p≤0.01), and with symptoms of at least mild depression or at least mild anxiety (PR:1.43, 95% CI 1.22-1.67, p≤0.01) (Table 2). In addition the prevalence of unmet need for contraception was higher among women who were younger (≤24 years) (PR:1.67, 95% CI 1.39–2.01, p≤0.01) and diagnosed with HIV within the past year (PR:1.60, 95% CI 1.27-2.00, p≤0.01), while being married or cohabiting (PR:0.37, 95% CI 0.27–0.50, p≤0.01), higher gravidity (≥3 vs 0-2; PR:0.56, 95% CI 0.44–0.73, p≤0.01), and disclosure of HIV status to a partner (PR:0.34, 95% CI 0.25–0.46, p≤0.01) were associated with a lower unmet need.

**Table 2:**
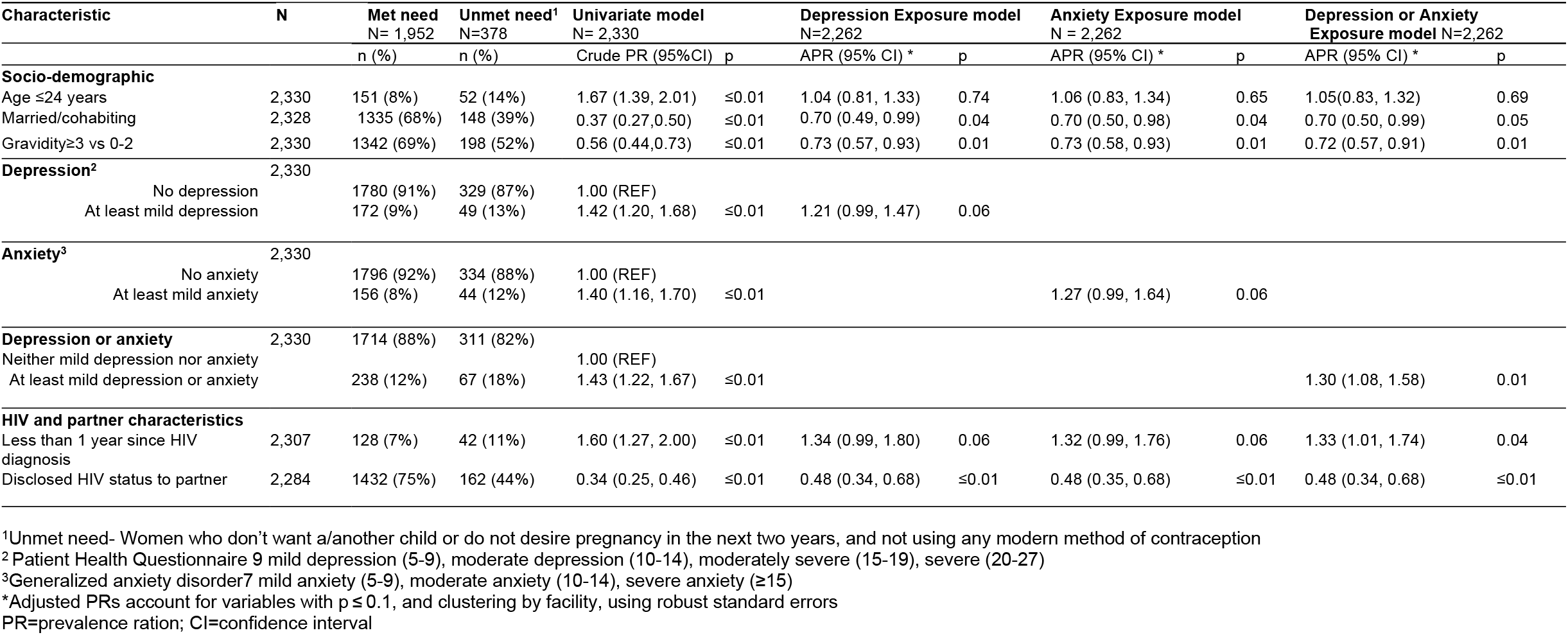
Relationship between depression, anxiety and other co-factors and unmet need for contraception among Kenyan women living with HIV.

| Characteristic | N | Met need<br>N= 1,952 | Unmet need<sup>1</sup><br>N=378 | Univariate model<br>N= 2,330 |  | Depression Exposure model<br>N=2,262 |  | Anxiety Exposure model<br>N = 2,262 |  | Depression or Anxiety<br>Exposure model N=2,262 |  |
| --- | --- | --- | --- | --- | --- | --- | --- | --- | --- | --- | --- |
|  |  | n (%) | n (%) | Crude PR (95%CI) | p | APR (95% CI) * | p | APR (95% CI) * | p | APR (95% CI) * | p |
| **Socio-demographic** |  |  |  |  |  |  |  |  |  |  |  |
| Age ≤24 years | 2,330 | 151 (8%) | 52 (14%) | 1.67 (1.39, 2.01) | ≤0.01 | 1.04 (0.81, 1.33) | 0.74 | 1.06 (0.83, 1.34) | 0.65 | 1.05(0.83, 1.32) | 0.69 |
| Married/cohabiting | 2,328 | 1335 (68%) | 148 (39%) | 0.37 (0.27,0.50) | ≤0.01 | 0.70 (0.49, 0.99) | 0.04 | 0.70 (0.50, 0.98) | 0.04 | 0.70 (0.50, 0.99) | 0.05 |
| Gravidity≥3 vs 0-2 | 2,330 | 1342 (69%) | 198 (52%) | 0.56 (0.44,0.73) | ≤0.01 | 0.73 (0.57, 0.93) | 0.01 | 0.73 (0.58, 0.93) | 0.01 | 0.72 (0.57, 0.91) | 0.01 |
| **Depression<sup>2</sup>** |  |  |  |  |  |  |  |  |  |  |  |
| No depression | 2,330 | 1780 (91%) | 329 (87%) | 1.00 (REF) |  |  |  |  |  |  |  |
| At least mild depression |  | 172 (9%) | 49 (13%) | 1.42 (1.20, 1.68) | ≤0.01 | 1.21 (0.99, 1.47) | 0.06 |  |  |  |  |
| **Anxiety<sup>3</sup>** |  |  |  |  |  |  |  |  |  |  |  |
| No anxiety | 2,330 | 1796 (92%) | 334 (88%) | 1.00 (REF) |  |  |  |  |  |  |  |
| At least mild anxiety |  | 156 (8%) | 44 (12%) | 1.40 (1.16, 1.70) | ≤0.01 |  |  | 1.27 (0.99, 1.64) | 0.06 |  |  |
| **Depression or anxiety** |  |  |  |  |  |  |  |  |  |  |  |
| Neither mild depression nor anxiety | 2,330 | 1714 (88%) | 311 (82%) | 1.00 (REF) |  |  |  |  |  |  |  |
| At least mild depression or anxiety |  | 238 (12%) | 67 (18%) | 1.43 (1.22, 1.67) | ≤0.01 |  |  |  |  | 1.30 (1.08, 1.58) | 0.01 |
| **HIV and partner characteristics** |  |  |  |  |  |  |  |  |  |  |  |
| Less than 1 year since HIV diagnosis | 2,307 | 128 (7%) | 42 (11%) | 1.60 (1.27, 2.00) | ≤0.01 | 1.34 (0.99, 1.80) | 0.06 | 1.32 (0.99, 1.76) | 0.06 | 1.33 (1.01, 1.74) | 0.04 |
| Disclosed HIV status to partner | 2,284 | 1432 (75%) | 162 (44%) | 0.34 (0.25, 0.46) | ≤0.01 | 0.48 (0.34, 0.68) | ≤0.01 | 0.48 (0.35, 0.68) | ≤0.01 | 0.48 (0.34, 0.68) | ≤0.01 |

<sup>1</sup>Unmet need- Women who don't want a/another child or do not desire pregnancy in the next two years, and not using any modern method of contraception
<sup>2</sup> Patient Health Questionnaire 9 mild depression (5-9), moderate depression (10-14), moderately severe (15-19), severe (20-27)
<sup>3</sup>Generalized anxiety disorder7 mild anxiety (5-9), moderate anxiety (10-14), severe anxiety (≥15)
\*Adjusted PRs account for variables with p ≤ 0.1, and clustering by facility, using robust standard errors
PR=prevalence ration; CI=confidence interval

In multivariable models, the association between at least mild depression and need was attenuated when adjusted for age, marital status, gravidity, time since HIV diagnosis, HIV (adjusted prevalence ratio [aPR] 1.21, 95% CI 0.99–1.47, p=0.06). The association between at least mild anxiety and unmet need was also attenuated after multivariable adjustment (aPR 1.27, 95% CI 0.99–1.64, p=0.06), while having at least mild depression or at least mild anxiety remained significantly associated with unmet need in the multivariable model (aPR 1.30, 95% CI 1.08–1.58, p=0.01).

In sensitivity analyses, we did not detect a significant relationship between having at least moderate depression or anxiety or having at least severe depression or anxiety and unmet need (Supplementary Table A).

### LARC use

Among modern contraceptive users (excluding permanent methods), LARC use was 41% (n=784). In univariate analyses, we did not detect a relationship between having at least mild depressive symptoms, at least mild anxiety or at least mild symptoms of depression or at least mild anxiety and LARC use (Table 3). However, LARC use was 37% lower among women who experienced stigma (PR:0.63, 95% CI 0.50–0.81, p≤0.01) than women without stigma. Findings from multivariable models examining LARC use were similar to univariate findings and stigma remained independently associated with lower LARC use across all three models.

**Table 3:** Relationship between depression, anxiety and other co-factors and LARC use among Kenyan women living with HIV.

| Characteristic | N | LARC<sup>1</sup><br>N= 784 | Short-acting<sup>2</sup><br>N=1,127 | Univariate model<br>N= 1,911 |  | Depression Exposure model<br>N=1,909 |  | Anxiety Exposure model<br>N= 1,909 |  | Depression or Anxiety<br>Exposure model N=1,909 |  |
| --- | --- | --- | --- | --- | --- | --- | --- | --- | --- | --- | --- |
|  |  | n (%) | n (%) | Crude PR (95%CI) | p | APR (95% CI)* | p | APR (95% CI)* | p | APR (95% CI)* | p |
| **Socio-demographic** |  |  |  |  |  |  |  |  |  |  |  |
| Age ≤ 24 years | 1,911 | 68 (9%) | 83 (7%) | 1.11 (0.89, 1.38) | 0.37 |  |  |  |  |  |  |
| Married/cohabiting | 1,910 | 558 (71%) | 748 (66%) | 1.15 (0.99, 1.32) | 0.06 | 1.08 (0.93, 1.24) | 0.31 | 1.08 (0.93, 1.24) | 0.31 | 1.08 (0.93, 1.24) | 0.31 |
| **Depression<sup>3</sup>** |  |  |  |  |  |  |  |  |  |  |  |
| No depression | 1,911 | 721 (92%) | 1023 (91%) | 1.00 (REF) |  |  |  |  |  |  |  |
| At least mild depression |  | 63 (8%) | 104 (9%) | 0.91 (0.73, 1.14) | 0.42 | 1.01 (0.87, 1.17) | 0.94 |  |  |  |  |
| **Anxiety<sup>4</sup>** |  |  |  |  |  |  |  |  |  |  |  |
| No anxiety | 1,911 | 723 (92%) | 1033 (92%) | 1.00 (REF) |  |  |  |  |  |  |  |
| At least mild anxiety |  | 61 (8%) | 94 (8%) | 0.96 (0.70, 1.31) | 0.78 |  |  | 1.06 (0.91, 1.23) | 0.45 |  |  |
| **Depression or anxiety** |  |  |  |  |  |  |  |  |  |  |  |
| Neither mild depression nor anxiety | 1,911 | 694 (89%) | 984 (87%) | 1.00 (REF) |  |  |  |  |  |  |  |
| At least mild depression or anxiety |  | 90 (11%) | 143 (13%) | 0.93 (0.73, 1.19) | 0.58 |  |  |  |  | 1.04 (0.93, 1.18) | 0.48 |
| **Stigma<sup>5</sup>** |  |  |  |  |  |  |  |  |  |  |  |
| Experienced stigma | 1,910 | 553 (71%) | 957 (85%) | 0.63 (0.50, 0.81) | ≤0.01 | 0.77 (0.65, 0.90) | ≤0.01 | 0.76 (0.65, 0.90) | ≤0.01 | 0.76 (0.65, 0.90) | ≤0.01 |

<sup>1</sup>LARC-long-acting reversible contraception; IUCD (intra-uterine contraceptive device) and Implant
<sup>2</sup>Short-acting- injectables, oral contraception, condoms, lactation amenorrhoea, vaginal rings, e-pills, standard days
<sup>3</sup>Patient Health Questionnaire 9 mild depression (5-9), moderate depression (10-14), moderately severe (15-19), severe (20-27)
<sup>4</sup>Generalized anxiety disorder 7 mild anxiety (5-9), moderate anxiety (10-14), severe anxiety (≥15)
<sup>5</sup>Berger HIV Stigma Scale- low (score 13-24), moderate (score 25-36), high (score 37-48)
\*Adjusted PRs account for variables with p ≤ 0.1, and clustering by facility, using robust standard errors
PR=prevalence ration; CI=confidence interval

Sensitivity analyses of having at least moderate depression or anxiety and LARC use showed similar findings as univariate models (Supplementary Table B).

## DISCUSSION

In this cohort of Kenyan WLHIV without pregnancy desires, we found that 16% of women had an unmet need for contraception. This estimate is similar to national population-level estimates for Kenya which report unmet need of 14% among women of reproductive age(29)(30). These data suggest barriers to contraceptive access or uptake are similar among WLHIV and the general population of women who wish to avoid pregnancy.

The prevalence of at least mild depression and at least mild anxiety were both, individually, 9%: collectively, 13% of women had either at least mild depression or anxiety. Our results were similar to another study among WLHIV in Kenya(31) which reported (~9%) of pregnant WLHIV reported depressive symptoms. In contrast, studies from other African settings have reported substantially higher prevalence of depressive and anxiety symptoms among WLHIV, including prevalences of 29% and 33% for anxiety and depression, respectively, in Ethiopia(32) and regional meta-analyses a pooled prevalence of depression of 38% among people living with HIV in East Africa(2). These results highlighting substantial regional variability.

We found that women who had at least mild depression or anxiety had a 30% higher unmet need which was a statistically significant association. While point estimates were similar for analysis examining depression alone or anxiety alone and unmet need, these results were of borderline significance, which suggests we may have lacked power to detect associations with individual condition symptomatology This may also explain why we did not observe associations between having at least moderate anxiety and/or depression: sample sizes of women with more severe symptoms were insufficient to detect this magnitude of association. There can be multiple potential explanations for why women who do not desire pregnancy, and also experience depression, may be less likely to use contraception. Depression, characterized by persistently low mood, energy and motivation, can negatively impact the likelihood of seeking care(11) or obtaining information about contraceptive options. Additionally, cognitive impairments associated with depression, such as difficulty with decision-making and prioritization, may prevent women from addressing their sexual and reproductive health needs, particularly when faced with concurrent stressors such as managing a recent HIV diagnosis. Our findings are consistent with prior research demonstrating how psychological distress can impair women’s ability to proactively seek contraceptive services, initiate discussions with providers, and maintain consistent use(33)(34). Major depression has previously been shown to be associated with elevated risks of both contraceptive non-use and inconsistent use(35) in a cohort of women in the US. An additional, another study conducted in the US, found that adolescents and young women with depressive symptoms, or who reported either anxiety or stress, were more likely to delay initiation of contraception.

While we did not detect a relationship between depression or anxiety and LARC, we did find that women who perceive stigma were significantly less likely to use LARC. Stigma, including within the healthcare setting, has been negatively associated with general delays in seeking care and negative health outcomes(36)(37) and specifically lower rates of contraceptive uptake, particularly for provider-dependent methods such as LARC(38)(34). Since LARC requires clinic attendance and direct provider involvement, stigma may present an important barrier to uptake of these methods(39). Stigma adds another layer to accessing LARC, as women with symptoms of depression or anxiety may not have the ability or self-efficacy to overcome these existing challenges.

In addition, we found additional correlates of unmet need. Unmet need for contraception was higher among younger women in univariate analysis; however, this relationship was no longer statistically significant in multivariate models. These results mirror trends observed in the general population in Kenya, where unmet need is 14% among all women and 25% among younger women aged 15-24 years(40)(41)(42). Higher unmet need among younger women may stem from limited knowledge, sociocultural constraints, and unequal power dynamics in relationships, as well as provider biases and poor privacy in service delivery(43)(42). Unmet need was higher among women diagnosed with HIV within the last year but lower among women who disclosed their HIV status to their partner. These results concur with findings from South African, where women who initiated ART within the past year were 50% more likely to be non-users of modern contraception (44). Findings from qualitative research in Zimbabwe also found that women newly diagnosed with HIV may struggle to prioritize reproductive health as they adjust to their diagnosis, navigate disclosure, and manage psychosocial distress(45).

Our study had several strengths but was also subject to some limitations. Screening for depression and anxiety using validated instruments provided an opportunity to assess symptoms strongly predictive of depression and anxiety conditions, an approach that can be used where resources are limited and help direct people who screen positive for additional assessments and care. Our study was conducted in multiple sites with a large sample of WLHIV without pregnancy intention, it was conducted in high HIV burden settings and our findings may not be generalizable to other contexts or settings, including those of lower HIV burden. Results are also subject to selection bias due to trial eligibility criteria, particularly mobile phone access. In addition, the cross-sectional study design limits assessment of the temporal relationship between exposure (depression and anxiety) and outcome (unmet need and method type).

In conclusion, our study contributes to the understanding of the relationship between depression and anxiety and unmet need for contraception among WLHIV in sub-Saharan Africa, including exploring factors unique to WLHIV, such as disclosure and stigma. Our results suggest that women who experience mental health conditions may find it more difficult to use contraception, and women with perceived stigma may face additional barriers to using LARC. Future studies should examine the underlying reasons for unmet need for contraception among WLHIV with depression, anxiety or stigma and identify strategies to better support them, which may help them achieve their reproductive health goals.

## Data Availability

De-identified data are available from Kenyatta National Hospital upon reasonable request by researchers who meet the criteria for access, subject to ethics approval and a data-use agreement.

## Notes

**Conflicts of Interest:** The authors have no financial conflicts of interest to declare

**Funding:** Funding for the study is provided by NIH-Fogarty 5D43TW009580 to AKK and NIH-NICHD R01HD104551. Support is also provided by the Global Center for Integrated Health of Women, Adolescents, and Children (Global WACh) and the University of Washington / Fred Hutch Center for AIDS Research, NIH-NIAID AI027757

### Competing Interest Statement

The authors have declared no competing interest.

### Author Declarations

The study procedures were approved by the University of Washington Human Subjects Division and Kenyatta National Hospital-University of Nairobi Ethics Committee, (KNH-UoN ERC)

